# A Refined Lung Transplant Frailty Scale Incorporating Serum Albumin Is Associated With Risk of Mortality in Lung Transplant Candidates

**DOI:** 10.64898/2026.10.08.26365085

**Authors:** Anya Sriram, Jason D Christie, Michaela R Anderson, Luke A Benvenuto, Ying Gao, Selim M Arcasoy, Daniel Calabrese, Steven R Hays, Jasleen Kukreja, Aida Venado, Legna Betancourt, Paige Gambetta, Hunter Scott, Michelle Oyster, Derek Zaleski, Joe Adler, Laurel Kalman, Shreena Patel, Michael Tevald, Kenneth E Covinsky, Chiung-Yu Huang, John R Greenland, Patricia Katz, Jonathan P Singer

## Abstract

The Lung Transplant Frailty Scale (LT-FS) is a validated binary measure of frailty in lung transplant candidates. We refined the LT-FS Base and Body Composition measures by developing a three-category classification to improve clinical risk stratification and replacing serum C-reactive protein with albumin as a more clinically practical frailty biomarker.

We applied a 4-step measure development method to a multicenter prospective cohort study of lung transplant candidates. Low albumin (<3.5 g/dL) replaced high serum C-reactive protein (≥10 mg/L) as the LT-FS biomarker. Cutpoints balancing discrimination, risk prediction, and calibration defined “frail”, “pre-frail”, and “not frail” categories. We then evaluated these category associations with exercise capacity, patient-reported outcomes, and mortality using multivariate linear and Cox regression.

Among 419 participants, the refined LT-FS identified a graded increase in mortality risk across frailty categories. Pre-frailty and frailty by LT-FS Body Composition were associated with 2.9-fold (HR 2.9, 95% CI 1.2–6.9) and 7-fold (HR 6.9, 95% CI 2.6–18.2) higher risks of waitlist delisting or death, respectively. After transplant, pre-frailty and frailty by LT-FS Body Composition were associated with 2-fold (HR 2.1, 95% CI 1.2–3.8) and 3.6-fold (HR 3.6, 95% CI 1.8–7.3) higher mortality risk, respectively. Relationships with exercise capacity and disability were less consistent.

A refined Lung Transplant Frailty Scale incorporating serum albumin demonstrates strong associations with waitlist and post-transplant mortality. The addition of a pre-frail category better captures the continuum of frailty and improves clinical risk stratification of lung transplant candidates.

## Introduction

Frailty has emerged as a strong risk factor for poorer health-related quality of life (HRQL), increased peri and post-operative complications, and increased mortality before and after lung transplantation^1–6^. As a result, frailty assessment has become an important part of the transplant evaluation process^7^. Efforts to refine the measurement of frailty in lung transplant candidates are an important area of active work.

We previously developed the Lung Transplant Frailty Scale (LT-FS), a novel measure designed to quantify frailty in those with advanced lung disease undergoing evaluation for lung transplantation^8^. The LT-FS has three versions: one composed of simple-to-collect physical performance measures and serum C-reactive protein (CRP; a frailty biomarker of systemic inflammation^9^) called the LT-FS Base; one that adds a measure of skeletal muscle mass to the Base instrument called the LT-FS Body Composition; and one that adds research-grade biomarkers to the LT-FS Body Composition instrument. Each of these instruments exhibited superior predictive validity when compared to other commonly used measures of physical frailty^8^.

As with many measures, increased experience and real-world implementation of the LT-FS in a clinical setting identified opportunities for refinement. Firstly, we found that some patients exhibited large day-to-day variations in CRP levels, raising concerns about stability of scores. Secondly, the original LT-FS yields a dichotomous categorization of “frail” or “not frail”. In reality, rather than an all-or-nothing condition, frailty likely represents a continuum, ranging from robust to severely frail. Extensively validated frailty measures, such as the Short Physical Performance Battery (SPPB) and Fried Frailty Phenotype (FFP), can be evaluated as a continuous or ordinal measure which includes a pre-frail category^1,10,11^. Although large enough samples sizes are still lacking to risk-stratify across continuous LT-FS scores, an ordinal LT-FS instrument that categorizes patients into “frail”, “pre-frail”, and “not frail” groups could be clinically valuable by capturing meaningful gradations.

Therefore, in this study, we explored two hypotheses to address the need for improved day-to-day reliability and granularity of the LT-FS. We first hypothesized that serum albumin could replace CRP as a clinically readily available and stable biomarker of frailty. Albumin is a well-described biomarker of frailty and generally exhibits greater day-to-day stability than CRP^12–14^. Secondly, we hypothesized that we could create a 3-category ordinal version of the LT-FS to refine risk stratification and discrimination for lung transplant candidacy.

## Methods

### Study Design

We analyzed participants in the multicenter observational Lung Transplant Body Composition (LTBC) cohort study. This cohort and analyses comprised lung transplant candidates (age ≥18) enrolled between June 2017 and November 2021 at UCSF, Columbia University Medical Center, and the University of Pennsylvania. Vital status was collected through August 2025. Center Institutional Review Boards approved this study and participants provided written informed consent for participation. This article is available as a preprint.

### Development of the original LT-FS

Frailty assessments were performed during the evaluation process near the time of placement on the waitlist for transplant^8^. From this, we developed the three LT-FS instruments, LT-FS Base, LT-FS Body Composition, and LT-FS Biomarker, with each instrument incorporating additional measures of frailty. LT-FS Base includes measures of balance, grip strength, gait speed, and CRP. LT-FS Body Composition adds appendicular skeletal muscle mass. These first two instruments were deemed deployable in the clinical environment. The LT-FS Biomarker instrument incorporates research-grade biomarkers and was intended to advance future research efforts rather than be used clinically.

### Rationale for substituting albumin in place of CRP

In the first iteration of the LT-FS Base, three routinely available biomarkers (hemoglobin, serum albumin, and CRP) were considered as viable candidates for inclusion. Ultimately, we retained CRP based on the extant frailty biomarker literature and its univariate strength of association with waitlist death or delisting^15^. The large day-to-day variations in CRP observed in some patients during clinical implementation of the LT-FS and greater stability of albumin over short durations of time suggested that albumin could be considered as a substitute biomarker^14^. Additionally, our prior work demonstrated a strong univariate association of low serum albumin with waitlist death or delisting for becoming too ill^15^.

### Rationale for developing an ordinal frailty score

Conceptually and clinically, frailty is a continuum ranging from robust to severely frail. In our prior work with the LT-FS, the SPPB, and the FFP, we identified associations between these measures as continuous exposures and outcomes of interest, including mortality^1,2,15^. This has also been shown in studies of cumulative deficit frailty measures^16,17^. Until a larger evidence base is available, continuous scores may be challenging for clinicians to interpret. However, an ordinal LT-FS scale that captures an intermediate category between “frail” and “not frail” could better represent this continuum.

Given that our aim is to improve the clinical utility of the LT-FS, we focused on refining the Base and Body Composition instruments, recognizing that the LT-FS Biomarker instrument was not intended for clinical use.

### Frailty measurement

Frailty assessments, including gait speed, grip strength, balance, and body composition by bioelectrical impedance analysis (BIA) were performed near the time of placement on the waitlist for transplant. Assessments were repeated, as possible, every three months while participants were listed. Assessments most proximal to the time of transplant were used for this analysis.

Skeletal muscle mass (Appendicular Skeletal Muscle Index [ASMI]) and percent body fat were quantified by BIA using the InBody S10 (InBody USA, Cerritos, CA). Low muscle mass was defined as ASMI ≤7.3 kg/m^2^ for men, and ≤5.8 kg/m^2^ for women^18^.

We measured grip strength using a handheld dynamometer (Stoelting, Wood Dale, IL). For men, low grip strength was defined as ≤29kg for BMI ≤24kg/m^2^, ≤30kg for BMI 24.1-28 kg/m^2^, and ≤32kg for BMI >28 kg/m^2^ ^10^. For women, low grip strength was defined as ≤17 kg for BMI ≤23 kg/m^2^, ≤17.3 kg for BMI 23.1–26 kg/m^2^, ≤18 kg for BMI 26.1–29 kg/m^2^, and ≤21 kg for BMI >29 kg/m^2^ ^10^.

Serum albumin was extracted from the electronic medical record at the time of frailty assessment. Low albumin was defined as <3.5mg/dL^19^.

Baseline demographics and clinical variables were extracted from the electronic medical record at the time of frailty assessment. These variables included age, race, BMI, pulmonary diagnosis category used for Lung Allocation Score (LAS) calculation, hemoglobin, creatinine, lung function, six-minute walk distance (6MWD), and LAS at the time of frailty assessment^20^.

### Outcomes of interest

As in our prior reports, pre-transplant delisting for becoming too ill to safely undergo transplant or death before transplant were included in our waitlist delisting or death outcome^1,8,17,21^. The post-operative outcome was mortality up to 3-years after transplant. Additionally, pre-transplant waitlist delisting for becoming too ill or death before 3 years after transplant was used as a composite outcome.

Pre-transplant patient-reported physical functioning/disability was assessed with the Lung Transplant Valued Life Activities scale (LT-VLA), which ranges from 0-3 with a difference of 0.3 reflecting the minimally important difference (MID); higher scores reflect worse physical functioning/disability^22^. HRQL was assessed with the Medical Outcomes Study Short Form-12 Physical Component Summary Scale (SF12-PCS) which ranges from 0-100 with a difference of 5 reflecting the MID; higher scores reflect better HRQL^23^.

### Statistical Approach

The previously established LT-FS Base includes measures of balance, grip strength, gait speed, and high CRP and LT-FS Body Composition adds low ASMI (Supplemental Table 1). In this study, we analyzed the LT-FS with low albumin (<3.5mg/dL) as the frailty biomarker for both instruments, replacing high CRP (≥10 mg/L). As in the original LT-FS, in step 1, continuous frailty domain scores were calculated by multiplying each domain variable by its adjusted beta coefficient and summing the domains^8^.

Next, to define three frailty categories (“frail,” “pre-frail,” and “not frail”), we selected two cut points that balanced key performance outcomes while ensuring adequate sample sizes within each category. Conceptually, the anticipated distribution would follow a gradient in which the largest proportion of patients are classified as not frail, followed by pre-frail and frail patients. This expected pattern is consistent with the broader frailty literature. Since clinicians are likely to be most interested in a score that identifies patients at an increased risk for frailty-attributable poor outcomes, we sought to maximize discrimination while also balancing risk prediction and calibration. Cut points were selected using a hierarchical ranking approach. Macro-AUC (discrimination) was prioritized as the primary performance criterion, after which secondary performance metrics, including maximum rescaled R² (discrimination and calibration) and Brier score (accurate risk prediction) were used to rank the top 20 candidate scores. Cut points were selected from those satisfying the required group proportions, with preference given to those demonstrating the best overall performance^8,24^.

We examined associations of the refined LT-FS instrument using albumin with preoperative physical disability, 6MWD, and HRQL using were examined using linear regression. LT-FS associations with preoperative delisting for becoming too ill or death were evaluated using logistic regression, while the association with 3-year post-transplant mortality was evaluated using a Cox proportional hazards model with the date of transplantation as time zero. Participants who underwent transplant were followed until death or censoring at 3 years post-transplant. All models were adjusted for age, sex, race, diagnosis, and center based on their biologic plausibility as confounders and to control for center effect. Since frailty represents a continuum from robust to severely frail, we modeled LT-FS both as a continuous exposure and as a categorical variable, and evaluated its association with outcomes of interest. The proportional hazards assumption was assessed using Schoenfeld residuals. Kaplan-Meier methods were used to visualize the association of frailty categories with preoperative delisting or death as well as with post-transplant mortality. Analyses were performed using SAS version 9.4 (SAS Institute, Cary, NC) and STATA version 18.0 (StataCorp, College Station, TX). Statistical significance was set at 0.05

## Results

Among the 577 participants who were listed for transplant during the study period, 419 participants had measures of frailty and body composition and made up the cohort for this analysis (Figure 1). Of the 419 participants, 200 (47.7%) were female and the median age was 61 (interquartile range [IQR] 51-66) (Table 1). Most participants underwent transplant for pulmonary fibrosis (67.1%), followed by 19.1% for non-suppurative obstructive lung disease, predominantly chronic obstructive pulmonary disease (COPD); 7.2% for suppurative lung diseases, predominantly cystic fibrosis; and 6.7% for pulmonary arterial hypertension. 41 (9.8%) participants were delisted or died before transplant, 18 (5.2%) died within the first year after transplant, and 61 (14.6%) died within the first three years after transplant. No participants were lost to follow-up.

**Figure 1.**
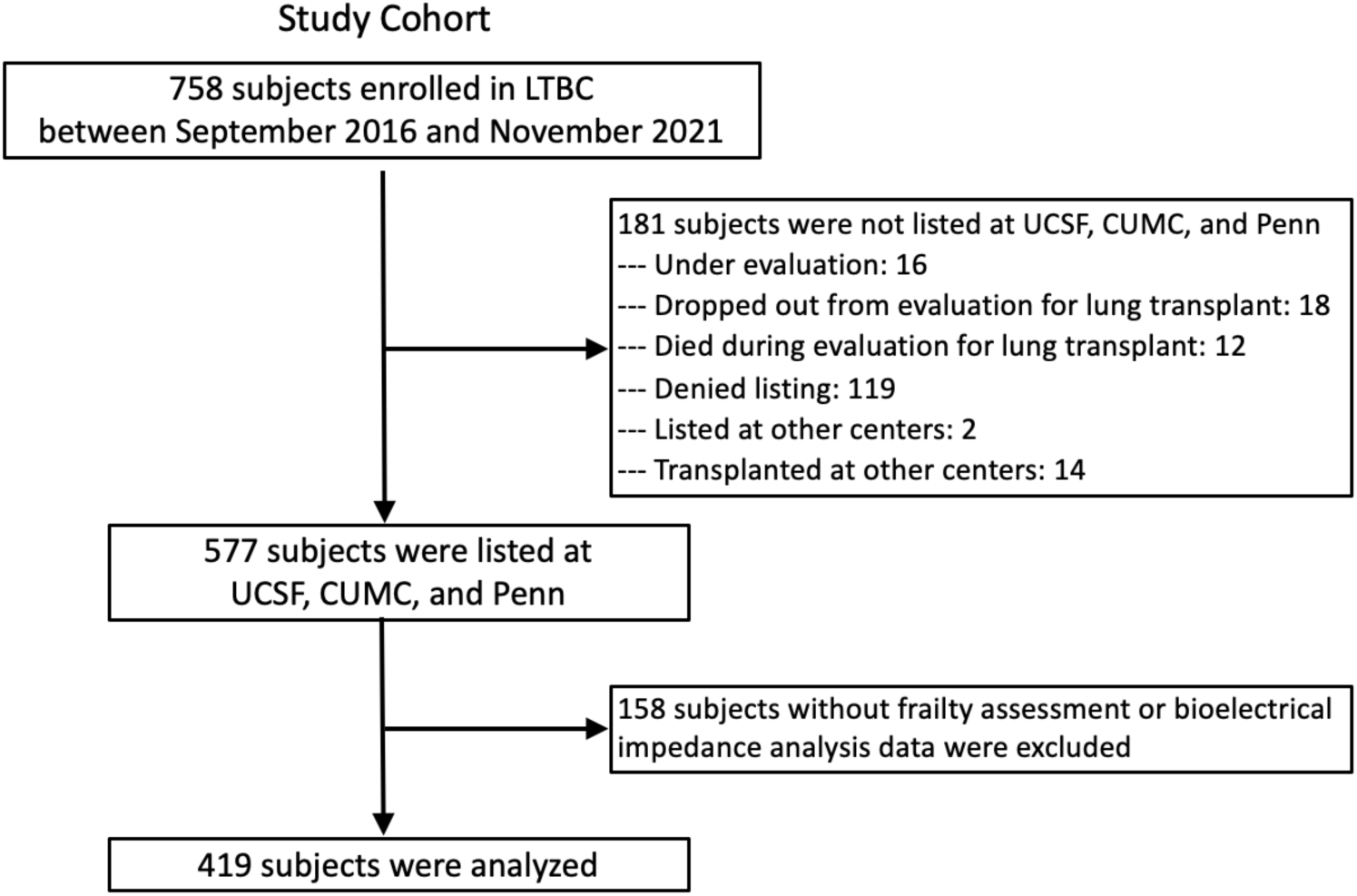
Study Flow Diagram

**Table 1.**
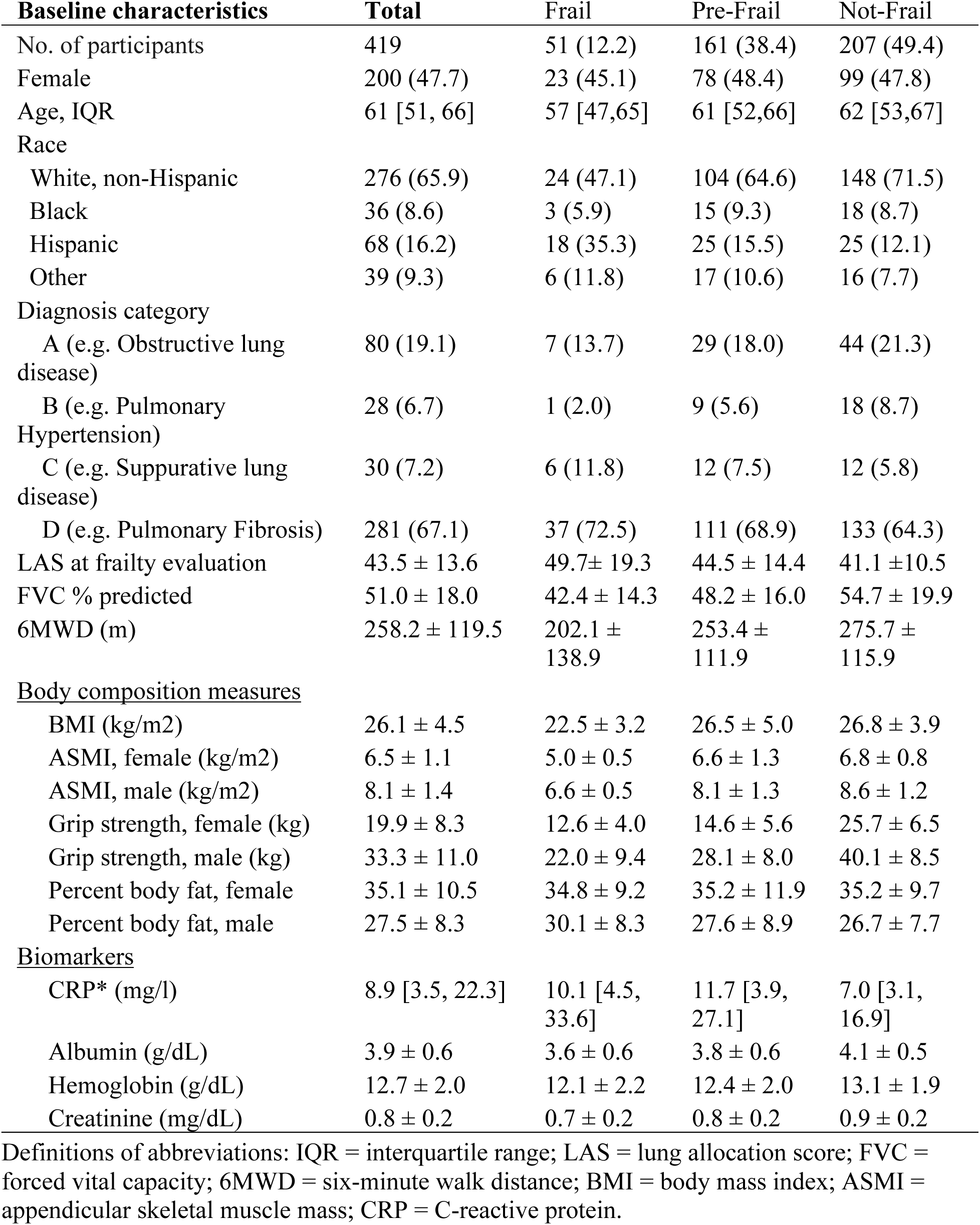
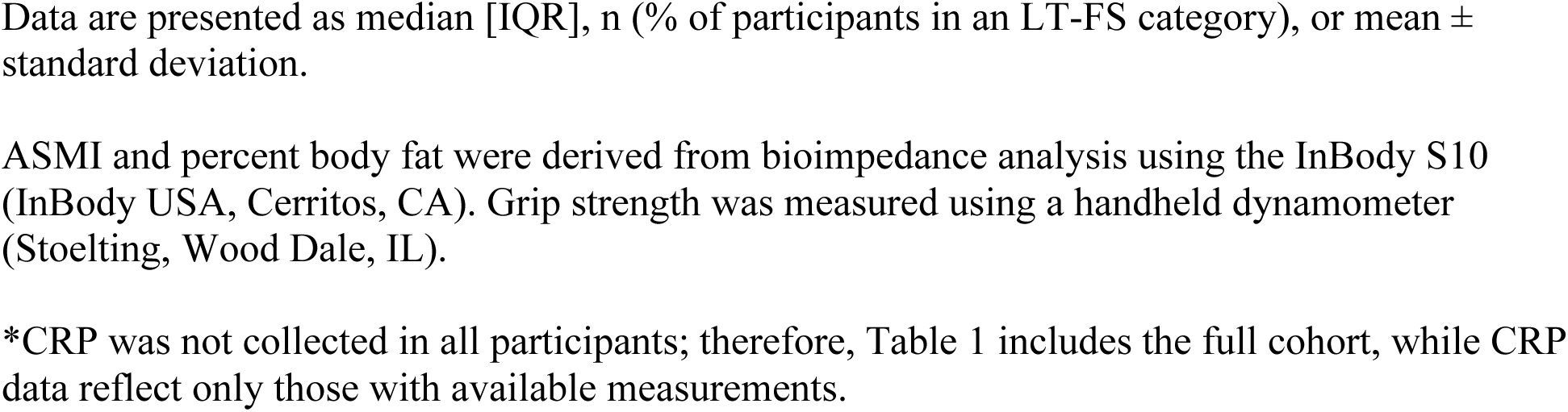
Demographics and Baseline Clinical Characteristics by LT-FS Body Composition defined categories. Definitions of abbreviations: IQR = interquartile range; LAS = lung allocation score; FVC = forced vital capacity; 6MWD = six-minute walk distance; BMI = body mass index; ASMI = appendicular skeletal muscle mass; CRP = C-reactive protein. Data are presented as median [IQR], n (% of participants in an LT-FS category), or mean ± standard deviation. ASMI and percent body fat were derived from bioimpedance analysis using the InBody S10 (InBody USA, Cerritos, CA). Grip strength was measured using a handheld dynamometer (Stoelting, Wood Dale, IL). *CRP was not collected in all participants; therefore, Table 1 includes the full cohort, while CRP data reflect only those with available measurements.

**Table 2.** Continuous refined LT-FS instrument characteristics and associations with mortality before and after transplant.

| **Instrument** | Mean ± SD | Score ranges | OR* (95% CI) per one point increase in score |
| --- | --- | --- | --- |
| **Base** | 14.5 ± 4.6 | 9.1 – 24.6 | 1.10 (1.05, 1.16)<br>p < 0.0001 |
| **Body Composition** | 15.6 ± 5.8 | 9 – 30.7 | 1.11 (1.06, 1.15)<br>p < 0.0001 |

\*OR calculated using composite outcome of waitlist delisting for becoming too ill or death and 3-year posttransplant mortality. Models adjusted for age, sex, and transplant center.
SD = standard deviation; OR = odds ratio; CI = confidence interval.

By the refined LT-FS Body Composition, 51 (12%) were frail, 161 (38%) were pre-frail, and 207 (49%) were not frail (Table 1). Non-frailty tended to be more prevalent in non-Hispanic white populations, and frailty tended to be more common in Hispanic and other, non-categorized, populations. Frailty tended to be more prevalent in those with suppurative lung disease and less common in those with pulmonary hypertension.

When analyzed as a continuous exposure, LT-FS was associated with increased risk of the composite outcome of waitlist delisting for becoming too ill or death or post-transplant mortality. For both LT-FS Base and Body Composition, each one-point worsening in frailty score was associated with a 10% increased risk of the composite outcome (Base OR: 1.1, 95% CI 1.05, 1.16; p<0.01; Body Composition OR: 1.1, 95% CI 1.06, 1.15; p<0.01).

When examining LT-FS Base and Body Composition as categorical variables, pre-frailty and frailty were associated with increased and progressive risk of mortality both before and after transplant (Table 3). Participants who were pre-frail or frail by LT-FS Base had a 2.5-fold and 8.3-fold increased risk of waitlist delisting for becoming too ill or death compared to non-frail participants, respectively (HR: 2.5, 95% CI 1.1,5.9; p = 0.03 for pre-frail; HR: 8.3, 95% CI 3.4, 20.5; p <0.01 for frail). And, for those who underwent transplant, pre-frailty and frailty by LT-FS Body Composition were associated with 2.1-fold and 3.6-fold increased risk of mortality up to 3-years after surgery, respectively (HR: 2.1, 95% CI 1.2, 3.8; p = 0.01 for pre-frail; HR: 3.6, 95% CI 1.8, 7.3; p <0.01 for frail). Of note, although pre-frailty by LT-FS Base was associated with a 1.7-fold increased risk of mortality after transplant, the association did not reach statistical significance (HR: 1.7, 95% CI: 0.9, 3.0; p = 0.10). Figure 2 shows Kaplan-Meier curves for waitlist delisting or death across the ordinal categories of the refined LT-FS Base and Body Composition instruments. Figure 3 shows post-transplant survival across the ordinal categories for both LT-FS instruments.

**Figure 2.**
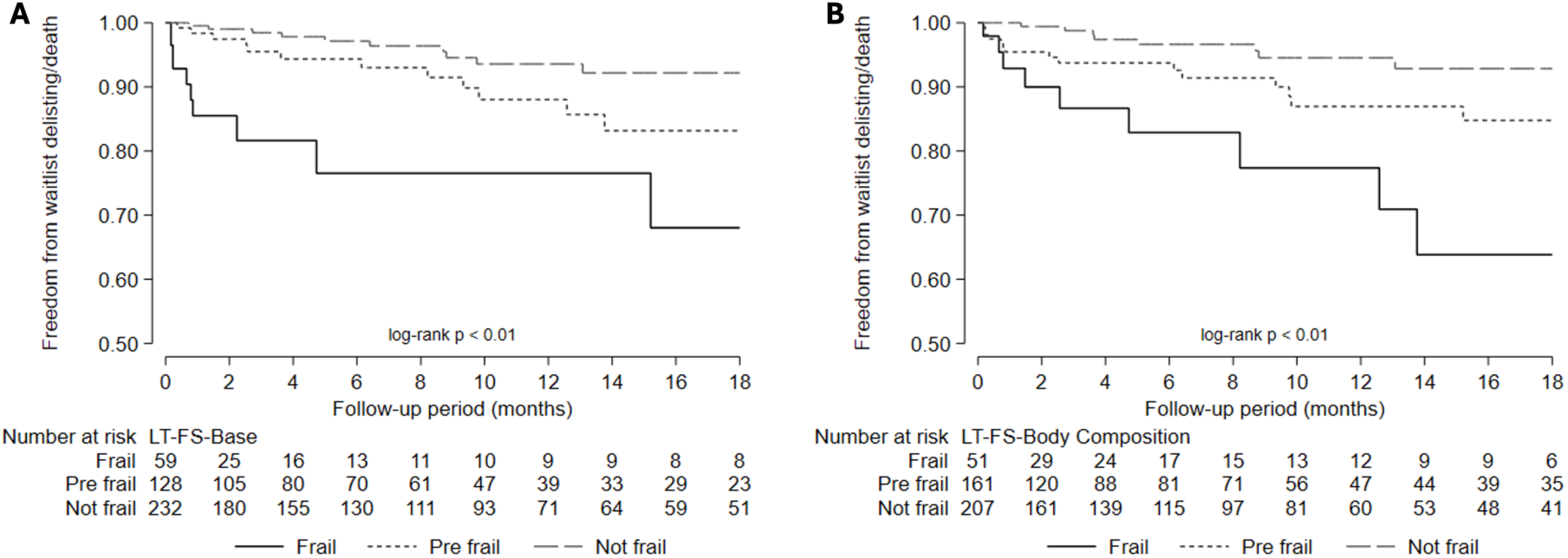
Association between LT-FS Base (A) and LT-FS Body Composition (B) categories and waitlist delisting for becoming too ill or death.

**Figure 3.**
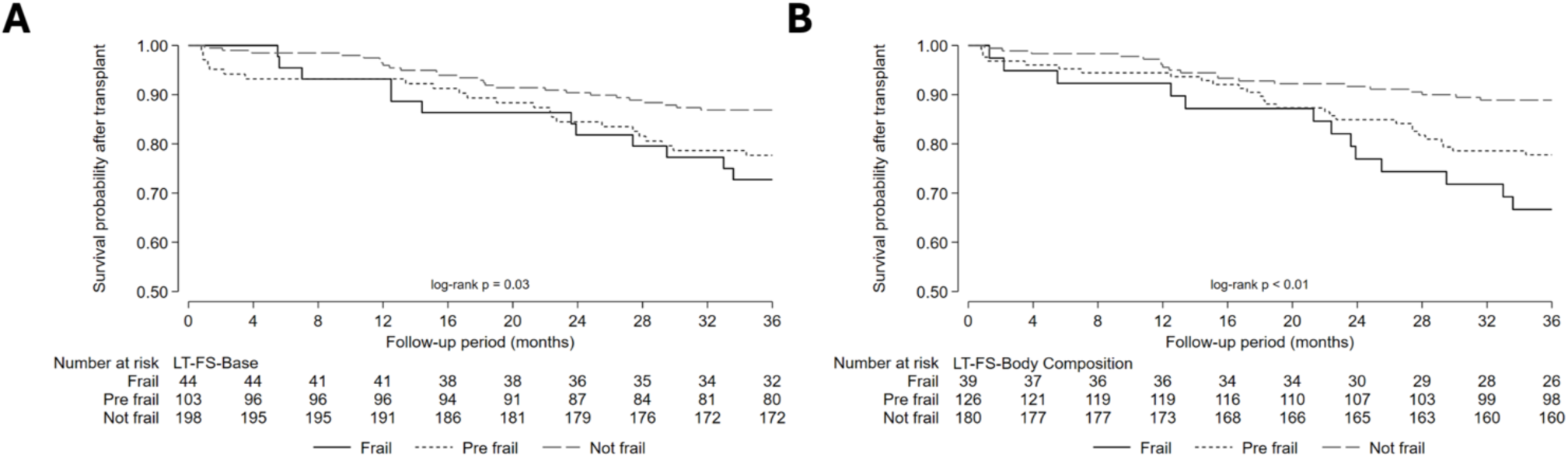
Association between LT-FS Base (A) and LT-FS Body Composition (B) categories and 3-year post transplant mortality.

**Table 3.** Risk of mortality by LT-FS frailty categories*.

| Instrument |  | Composite<br>outcome of<br>waitlist delisting<br>or death or death<br>after transplant<sup>†</sup> | Waitlist delisting<br>for becoming too<br>ill or death<sup>†</sup> | Post-transplant<br>mortality<sup>‡</sup> |
| --- | --- | --- | --- | --- |
| Base | Pre-Frail | 1.9 (1.1, 3.2)<br>p = 0.021 | 2.5 (1.1, 5.9)<br>p = 0.033 | 1.7 (0.9, 3.0)<br>p = 0.100 |
|  | Frail | 3.6 (1.9, 6.8)<br>p < 0.001 | 8.3 (3.4, 20.5)<br>p < 0.001 | 2.1 (1.1, 4.3)<br>p = 0.030 |
| Body<br>Composition | Pre-Frail | 2.5 (1.5, 4.3)<br>p < 0.001 | 2.91 (1.2, 6.9)<br>p = 0.016 | 2.1 (1.2, 3.8)<br>p = 0.014 |
|  | Frail | 5.9 (2.9, 11.7)<br>p < 0.001 | 6.9 (2.6, 18.2)<br>p < 0.001 | 3.6 (1.8, 7.3)<br>p < 0.001 |

\*Not frail is referent group. Models adjusted for age, sex, and transplant center. Post-transplant mortality through three-years.
<sup>†</sup>Data for the composite outcome are presented as Odds Ratios (95% confidence interval).
<sup>‡</sup>Data for waitlist delisting for becoming too ill or death and post-transplant mortality are presented as Hazard Ratios (95% confidence interval).

When examining physical functioning and disability by the LT-VLA, the association with pre-frailty and frailty differed between instruments. Neither pre-frailty by LT-FS Base or Body Composition nor frailty by LT-FS Body Composition were significantly associated worse LT-VLA scores. For example, LT-VLA scores were similar between those who were not frail and pre-frail by LT-FS Base and Body Composition (mean difference: 0.02, 95% CI −0.13, 0.17; p = 0.80 by Base, mean difference: 0.04, 95% CI −0.1, 0.2; p = 0.62 by Body Composition). Compared to non-frailty, frailty by LT-FS Base was associated with substantively worse patient-reported physical functioning (mean difference: 0.4, 95% CI: 0.17, 0.62; p <0.01).

Frailty by both LT-FS Base and Body Composition and pre-frailty by LT-FS Body Composition were associated with worse exercise capacity. Compared with participants who were not frail, those classed as frail by LT-FS Base and Body Composition had, on average, 111.4 meter (95% CI −145.5, −77.3; p <0.01) and 75.5 meter (95% CI −114.2, −36.9; p<0.01) shorter 6MWD, respectively. Pre-frailty was associated with a shorter 6MWD by LT-FS Body Composition (mean difference: −28.7m, 95% CI −53.0, −4.4; p = 0.02), but not by LT-FS Base (mean difference: −19.4m, 95% CI −45.0, 6.3; p = 0.14).

In the analysis of health-related quality of life by the SF12-PCS, the association with pre-frailty and frailty differed between instruments. Compared to non-frailty, pre-frailty by the Base and Body Composition instruments was not significantly associated with worse HRQL. Compared to those who were not frail, pre-frailty by Base and Body Composition was associated with an insignificant change in SF12-PCS score (mean difference: −0.4, 95% CI −2.7, 2.0; p = 0.74 by Base, mean difference: −0.7, 95% CI −2.9, 1.5; p = 0.52 by Body Composition). Frailty by LT-FS Base was associated with significantly worse HRQL (mean difference −3.9, 95% CI −7.4, −0.4; p = 0.03).

**Table 4.** Difference in HRQL, disability, and 6MWD by LT-FS frailty categories*.

| **Instrument** |  | **SF12-PCS** | **LT-VLA** | **6MWD (m)** |
| --- | --- | --- | --- | --- |
| Base | Pre-Frail | -0.4 (-2.7, 2.0)<br>p = 0.743 | 0.02 (-0.1, 0.2)<br>p = 0.798 | -19.4 (-45.0, 6.3)<br>p = 0.139 |
|  | Frail | -3.9 (-7.4, -0.4)<br>p = 0.03 | 0.4 (0.2, 0.6)<br>p = 0.001 | -111.4 (-145.5, -77.3)<br>p < 0.001 |
| Body Composition | Pre-Frail | -0.7 (-2.9, 1.5)<br>p = 0.522 | 0.04 (-0.1, 0.2)<br>p = 0.62 | -28.7 (-53.0, -4.4)<br>p = 0.021 |
|  | Frail | 0.6 (-3.1, 4.3)<br>p = 0.751 | 0.04 (-0.2, 0.3)<br>p = 0.771 | -75.5 (-114.2, -36.9)<br>p < 0.001 |

\*Not frail is referent group. Models adjusted for age, sex, and transplant center. SF12-PCS MID = 5, LT-VLA MID = 0.3.
HRQL = health-related quality of life; 6MWD = six-minute walk distance; LT-VLA = Lung Transplant Valued Life Activities; SF12-PCS = Short Form-12 Physical Component Summary Scale; MID = minimally important difference.
Data are presented as mean difference (95% confidence interval).

## Discussion

Using a multicenter cohort of adult lung transplant candidates, we refined the Lung Transplant Frailty Scale (LT-FS) by substituting serum albumin for CRP and by deriving a three-category scoring system (“not frail”, “pre-frail”, or “frail”). The refined LT-FS identifies graded mortality risk as both a continuous and ordinal measure. The continuous analysis—in which each one-point worsening in LT-FS was associated with a 10% increase in risk of mortality—supports that frailty is a continuum rather than an “all-or-nothing” state. The ordinal categorization captures the continuum with greater resolution than a dichotomous “frail versus not frail” classification, by identifying a pre-frail group that carries intermediate mortality risk before and after transplant.

The intermediate risk of mortality identified by the pre-frail category has direct clinical relevance. Notably, factors including BMI, 6MWD, and patient-reported functioning and HRQL were similar between pre-frail and not-frail lung transplant candidates. This suggests that pre-frailty is unlikely to be evident through routine clinical assessment alone. Recent literature has suggested that targeted interventions in the pre-operative period may improve post-operative outcomes^25,26^. In both the pre-frail and frail groups, interventions such as exercise and nutrition optimization or pulmonary rehabilitation could be employed to improve physiologic reserve prior to transplantation^25–28^.

In contrast to the strong association between the LT-FS and mortality, we found that associations between the LT-FS and HRQL or patient-reported disability were inconsistent. For example, frailty by LT-FS Base was associated with worse physical functioning, disability, and HRQL. Conversely, using the LT-FS Body Composition instrument, pre-frailty and frailty were not significantly associated with worse physical functioning or HRQL. Taken together, our findings suggest the mortality risk captured by the LT-FS is not reliably reflected in patient-reported functional status or HRQL.

The absence of a consistent association between LT-FS categories and HRQL differs from previous work in lung transplantation in which frailty was measured as a binary variable^4,8^. It is possible that use of a summary score for measuring HRQL obscured domain-level differences (e.g. physical function or social functioning) that vary by frailty category.

Our refinement to the LT-FS is consistent with other areas of clinical medicine in which syndrome definitions are iteratively updated as evidence accumulates and clinical experience identifies operational challenges. For example, in 2018, Thygesen et al. revised the universal definition of myocardial infarction in response to increasingly sensitive assays for myocardial injury^29^. Here, we updated a frailty measure specific to advanced lung disease and lung transplant patients by refining its biomarker and scoring structure to improve its clinical utility.

Serum albumin is a pragmatic choice for a frailty biomarker. It is influenced by nutritional status and systemic inflammation, and frailty is associated with low albumin across population studies^30,31^. Further, it is routinely collected during lung transplant candidacy evaluations, shows greater day-to-day stability than CRP, and hypoalbuminemia is independently associated with mortality after lung transplantation^14,19^.

Our study has limitations. Firstly, recruitment and data collection was limited by the COVID19 pandemic, which precluded a sample size large enough to enable a derivation and validation analysis. Thus, our findings should be considered preliminary until they can be validated in another population. Further, the study cohort that had available albumin values was substantially larger than the cohort with CRP. Thus, direct comparison of the albumin- and CRP-based LT-FS models could not be performed, and so the comparative performance between these two models remains undefined. Nonetheless, serum albumin is routinely collected as part of transplant evaluation and demonstrates greater day-to-day stability than CRP, supporting its utility as a clinically accessible frailty biomarker and use in this frailty measure. Additionally, we only included first time lung transplant candidates, which impeded our ability to consider risk stratification in re-transplant candidates. In the post-transplant setting, differences in rehabilitation and recovery trajectories between participants were not captured.

Despite these limitations, our study has important strengths. This study was derived from a prospective, multicenter cohort designed explicitly to refine the quantification of frailty in lung transplant candidates. The methodological strengths of the original LT-FS report are carried through here, including following a validated stepwise approach to deriving three frailty categories. The result is a more refined instrument that substitutes a clinically convenient biomarker and provides improved risk stratification.

In conclusion, the refined Lung Transplant Frailty Scale identifies a graded mortality risk before and after lung transplant. The discrepancy between mortality risk and patient-reported outcomes suggest that pre-frailty by LT-FS may identify physiologic vulnerability that precedes symptomatic burden. Further study of targeted interventions in candidates identified as frail or pre-frail by LT-FS may be warranted.

## Supporting information

Suppplement

## Data Availability

All data produced in the present study are available upon reasonable request to the authors

## Financial Disclosure

The authors declare the following financial interests/personal relationships which may be considered as potential competing interests: J.P.S. reports financial support was provided by the National Institutes of Health. J.P.S.: Consulting fees from XVIVO Perfusion; Chair DSMB: Krystal Biotech. M.R.A: clinical trial Principal Investigator: United Therapeutics. J.C.: Consulting fees from GSK and United Health Care; DSMB NIH PETALnet and NHLBI. J.R.G: Consulting fees from Sobi and Arda Therapeutics. The other authors declare that they have no known competing financial interests or personal relationships that could have appeared to influence the work reported in this paper.

## Notes

### Author Declarations

Ethics committee/IRB of University of California San Francisco gave ethical approval for this work. Ethics committee/IRB of Columbia University Medical Center gave ethical approval for this work. Ethics committee/IRB of University of Pennsylvania gave ethical approval for this work.

### Summary of Updates

Financial disclosure updated; author affiliations updated.

### Funding

- NHLBI: R01 HL134851, HL151552
- VA CDA: IK2CX002011, U01HL145435, K24HL115354

