## Supplementary material for "A Refined Lung Transplant Frailty Scale Incorporating Serum Albumin Is Associated With Risk of Mortality in Lung Transplant Candidates": Suppplement

Supplemental figures.

Supplemental Table 1. Component elements of the LT-FS and cutoff point

| **LT-FS Component** | Cut point |
|---|---|
| SPPB Balance | Side-by-side tandem for 10 seconds, semi-tandem for 10 seconds, but unable to stand full tandem for >3 seconds*[1] |
| SPPB Slow gait speed (s) | <6.2 |
| FFP Weak grip (kg) | |
| M, BMI ≤24 | ≤29 |
| M, BMI 24.1-28 | ≤30 |
| M, BMI >28 | ≤32 |
| F, BMI ≤23 | ≤17 |
| F, BMI 23.1-26 | ≤17.3 |
| F, BMI 26.1-29 | ≤18 |
| F, BMI >29 | ≤21 |
| Body Composition | |
| Low ASMI (kg/m$^2$) | M ≤7.3; F≤5.8 |
| Biomarkers | |
| Low albumin (g/dL) | <3.5 |
| High CRP (mg/l) | ≥10 |

SPPB = Short Physical Performance Battery; FFP = Fried Frailty Phenotype; BMI = body mass index; ASMI = appendicular skeletal muscle mass; CRP = C-reactive protein.

ASMI was derived from bioimpedance analysis using the InBody S10. Grip strength was measured using a handheld dynamometer. SPPB gait speed conducted over a 4m distance.

*Patients who fail an easier balance test do not advance; thus, a cutoff of <3 points corresponds to completing side-by-side and semi-tandem (1 point each) but failing or holding the full tandem for <3 seconds (0 points).

Supplemental Table 2. Demographics and Baseline Clinical Characteristics by LT-FS Base defined categories.

| **Baseline characteristics** | **Total** | Frail | Pre-Frail | Not-Frail |
|---|---|---|---|---|
| No. of participants | 419 | 59 | 128 | 232 |
| Female | 200 (47.7) | 28 (47.5) | 60 (46.9) | 112 (48.3) |
| Age, IQR | 61 [51, 66] | 58 [47,65] | 63 [55,68] | 61 [50,66] |
| Race | | | | |
| White, non-Hispanic | 276 (65.9) | 27 (45.8) | 92 (71.9) | 157 (67.7) |
| Black | 36 (8.6) | 10 (16.9) | 8 (6.3) | 18 (7.8) |
| Hispanic | 68 (16.2) | 17 (28.8) | 20 (15.6) | 31 (13.4) |
| Other | 39 (9.3) | 5 (8.5) | 8 (6.3) | 26 (11.2) |
| Diagnosis category | | | | |
| A (e.g. Obstructive lung disease) | 80 (17.5) | 7 (11.9) | 18 (14.1) | 55 (23.7) |
| B (e.g. Pulmonary Hypertension) | 28 (6.7) | 0 (0.0) | 9 (7.0) | 19 (8.2) |
| C (e.g. Suppurative lung disease) | 30 (7.0) | 6 (10.2) | 8 (6.3) | 16 (6.9) |
| D (e.g. Pulmonary Fibrosis) | 281 (68.7) | 46 (78.0) | 93 (72.7) | 142 (61.2) |
| LAS at frailty evaluation | 43.5 ± 13.6 | 54.5 ± 19.6 | 43.0 ± 13.3 | 40.9 ± 10.3 |
| FVC % predicted | 51.0 ± 18.0 | 39.0 ± 13.6 | 50.3 ± 15.1 | 53.9 ± 19.8 |
| 6MWD (m) | 258.2 ± 119.5 | 166.0 ± 119.9 | 266.9 ± 112.1 | 276.8 ± 112.9 |
| Body composition measures | | | | |
| BMI (kg/m2) | 26.1 ± 4.5 | 26.1 ± 4.5 | 26.4 ± 4.8 | 26.0 ± 4.4 |
| ASMI, female (kg/m2) | 6.5 ± 1.1 | 6.2 ± 1.2 | 6.6 ± 1.4 | 6.6 ± 1.0 |
| ASMI, male (kg/m2) | 8.1 ± 1.4 | 7.6 ± 1.2 | 8.0 ± 1.4 | 8.4 ± 1.3 |
| Grip strength, female (kg) | 19.9 ± 8.3 | 11.9 ± 5.3 | 13.6 ± 4.6 | 25.2 ± 6.2 |
| Grip strength, male (kg) | 33.3 ± 11.0 | 21.3 ± 8.9 | 26.7 ± 6.4 | 40.1 ± 8.3 |
| Percent body fat, female | 35.1 ± 10.5 | 37.5 ± 10.3 | 34.9 ± 12.1 | 34.7 ± 9.6 |
| Percent body fat, male | 27.5 ± 8.3 | 30.0 ± 10.1 | 28.6 ± 7.4 | 26.2 ± 8.0 |
| Biomarkers | | | | |
| CRP* (mg/l) | 8.9 [3.5,22.3] | 22.8 [9.1,46.8] | 18.9 [3.8,9.4] | 7.7 [3.1,17.7] |
| Albumin (g/dL) | 3.9 ± 0.6 | 3.3 ± 0.7 | 4.0 ± 0.4 | 4.0 ± 0.5 |
| Hemoglobin (g/dL) | 12.7 ± 2.0 | 11.4 ± 2.1 | 13.0 ± 1.7 | 12.9 ± 2.0 |
| Creatinine (mg/dL) | 0.8 ± 0.2 | 0.7 ± 0.3 | 0.8 ± 0.2 | 0.8 ± 0.2 |

Definitions of abbreviations: IQR = interquartile range; LAS = lung allocation score; FVC = forced vital capacity; 6MWD = six-minute walk distance; BMI = body mass index; ASMI = appendicular skeletal muscle mass; CRP = C-reactive protein.

Data are presented as median [IQR], n (% of participants in an LT-FS category), or mean ± standard deviation.

ASMI and percent body fat were derived from bioimpedance analysis using the InBody S10 (InBody USA, Cerritos, CA). Grip strength was measured using a handheld dynamometer (Stoelting, Wood Dale, IL).

*CRP was not collected in all participants; therefore, Table 1 includes the full cohort, while CRP data reflect only those with available measurements.

Supplemental Table 3. Equations for Refined LT-FS using Serum Albumin.

| | | Definitions of scoring categories | | |
|---|---|---|---|---|
| **Instrument** | **Score equation** | Frail | Pre-Frail | Not Frail |
| Base | (((SPPB_Balance * (-0.0865)) +(FFP_Weak_Grip * 0.7851) +(SPPB_Slow_Gait_Speed * 0.2322) + (low_albumin * 0.4451)) + 1)* 10 | >19.3 | ≥15.9 and ≤19.3 | <15.9 |
| Body Composition | (((SPPB_Balance * (-0.1006)) + (FFP_Weak_Grip * 0.7203) + (SPPB_Slow_Gait_Speed *0.1318) + (low_albumin *0.3297) + (low_ASMI * 0.8888)) + 1) * 10 | >22.2 | ≥17.2 and ≤22.2 | <17.2 |

If a cut point is reached, the value assigned to the corresponding variable is 1, and if it is not reached, the value assigned is 0. For example, if a male patient with a BMI of 20 $kg/m^2$ had a grip strength of 20 kg, FFP_Weak_Grip would be assigned a value of 1.

Supplemental Table 4. LT-FS component variable names

| **LT-FS Component** | Variable Name |
|---|---|
| SPPB Balance | SPPB_Balance |
| SPPB Slow gait speed (s) | SPPB_Slow_Gait_Speed |
| FFP Weak grip (kg) | |
| M, BMI ≤24<br>M, BMI 24.1-28<br>M, BMI >28<br>F, BMI ≤23<br>F, BMI 23.1-26<br>F, BMI 26.1-29<br>F, BMI >29 | FFP_Weak_Grip |
| Body Composition | |
| Low ASMI ($kg/m^2$) | low_ASMI |
| Biomarkers | |
| Low albumin (g/dL) | low_albumin |
